# KidneyNetwork: Using kidney-derived gene expression data to predict and prioritize novel genes involved in kidney disease

**DOI:** 10.1101/2021.03.10.21253054

**Authors:** Floranne Boulogne, Laura R. Claus, Henry Wiersma, Roy Oelen, Floor Schukking, Niek de Klein, Shuang Li, Harm-Jan Westra, Bert van der Zwaag, Franka van Reekum, Dana Sierks, Ria Schönauer, Jan Halbritter, Nine V.A.M. Knoers, Genomics England Research Consortium, Patrick Deelen, Lude Franke, Albertien M. van Eerde

## Abstract

**Background:** Genetic testing in patients with suspected hereditary kidney disease may not reveal the genetic cause for the disorder as potentially pathogenic variants can reside in genes that are not yet known to be involved in kidney disease. To help identify these genes, we have developed KidneyNetwork, that utilizes tissue-specific expression to predict kidney-specific gene functions.

**Methods:** KidneyNetwork is a co-expression network built upon a combination of 878 kidney RNA-sequencing samples and a multi-tissue dataset of 31,499 samples. It uses expression patterns to predict which genes have a kidney-related function and which (disease) phenotypes might result from variants in these genes. We applied KidneyNetwork to prioritize rare variants in exome sequencing data from 13 kidney disease patients without a genetic diagnosis.

**Results:** KidneyNetwork can accurately predict kidney-specific gene functions and (kidney disease) phenotypes for disease-associated genes. Applying it to exome sequencing data of kidney disease patients allowed us to identify a promising candidate gene for kidney and liver cysts: *ALG6*.

**Conclusion:** We present KidneyNetwork, a kidney-specific co-expression network that accurately predicts which genes have kidney-specific functions and can result in kidney disease. We show the added value of KidneyNetwork by applying it to kidney disease patients without a molecular diagnosis and consequently, we propose *ALG6* as candidate gene in one of these patients. KidneyNetwork can be applied to clinically unsolved kidney disease cases, but it can also be used by researchers to gain insight into individual genes in order to better understand kidney physiology and pathophysiology.

**Significance statement:** Genetic testing in patients with suspected hereditary kidney disease may not reveal the genetic cause for the patient’s disorder. Potentially pathogenic variants can reside in genes not yet known to be involved in kidney disease, making it difficult to interpret the relevance of these variants. This reveals a clear need for methods to predict the phenotypic consequences of genetic variation in an unbiased manner. Here we describe KidneyNetwork, a tool that utilizes tissue-specific expression to predict kidney-specific gene functions. Applying KidneyNetwork to a group of undiagnosed cases identified *ALG6* as a candidate gene in cystic kidney and liver disease. In summary, KidneyNetwork can aid the interpretation of genetic variants and can therefore be of value in translational nephrogenetics and help improve the diagnostic yield in kidney disease patients.

## Introduction

Genetic testing in patients with suspected hereditary kidney disease can reveal causative pathogenic variants in kidney-related genes. However, in many cases, a genetic cause cannot yet be detected. Pathogenic variants in known kidney-related genes are detected in approximately 10-30% of genetically tested patients with chronic kidney disease of any cause^1–3^. However, these percentages are likely underestimations of the number of patients with a monogenic cause as variants in genes not yet implicated in kidney disease will go unnoticed. Potentially harmful variants can reside in these genes, which makes it difficult to prioritize and interpret the relevance of these variants. Therefore, in the current era of genomic medicine, one of the main challenges after a negative diagnostic result in known genes is to detect and prioritize new candidate genes with potentially pathogenic variants that can explain the patient’s disease.^4^

Several tools have been developed to predict candidate disease genes using RNA-sequencing data^5^. We recently developed GeneNetwork and the GeneNetwork-Assisted Diagnostic Optimization (GADO) method to prioritize new candidate disease genes based on RNA-sequencing data^6^. The idea behind this method is that certain rare disorders can be caused by variants in several genes. While these genes are different, they usually have similar biological functions. When studying gene expression data from a large number of samples, these disease genes usually show strong co-expression. Thus, if there are other genes that are strongly co-expressed with known rare disease genes, it is possible that variants in these other genes can also cause the same disease.

For this kind of tool to work optimally, the co-expression information should be as accurate as possible. For GADO, we built a gene co-expression network based on publicly available RNA-sequencing datasets from many different tissues and used this network to predict which genes might be causing rare diseases. These predictions were trained using the human phenotype ontology (HPO) database^7^. In the HPO database, genes are assigned to phenotypes − called HPO terms − that are based on gene−disease annotations and disease symptoms present in the OMIM^8^ and Orphanet^9^ databases. By integrating the information from the HPO database with the gene co-expression network, we could calculate prediction scores for each gene per HPO term. Together, these scores constitute GeneNetwork. The GADO method was then developed to identify genes with putative causal variants in patients with a (suspected) monogenic disease. GADO prioritizes genes by combining an input list of HPO terms that describe the patient’s phenotype with a list of genes with possible deleterious variants from that patient. The prioritization of the gene list is based on the combined gene prediction scores for the input HPO terms^6^.

Because we observed that GeneNetwork’s prediction performance for kidney-related phenotypes was limited, we sought to improve prediction by developing a kidney-specific network that uses a combination of GeneNetwork and 878 kidney RNA-sequencing samples. We also further improved the underlying prediction algorithms of the GADO method. In this paper we present the resulting KidneyNetwork, a co-expression network that can be used to accurately predict gene−phenotype associations of genes unknown for kidney-related HPO terms. As a proof of principle, we applied KidneyNetwork to exome sequencing data from a group of patients with previously unresolved kidney diseases.

## Methods

To improve the prediction of kidney-related phenotypes, we collected kidney-derived RNA-sequencing data, updated GeneNetwork with more recent reference databases and improved statistical analyses, followed by integration of tissue-specific information.

### Data collection for building KidneyNetwork

We built KidneyNetwork by combining an existing multi-tissue RNA-sequencing dataset with RNA-sequencing data from kidney samples to enhance kidney-specific signals. The selected kidney samples were from several origins, including primary, tumor and fetal tissue. We chose to include the multi-tissue dataset for two reasons. First, we needed a sufficient number of samples to build a baseline network. Second, we wanted to preserve expression that is specific to several, or all, kidney cell types but not to other tissues. We did this because gene−phenotype scores are based on differences in expression between samples; if all genes have high (or low) expression in all the samples included in the analysis, they will not add sufficient information to the prediction algorithm.

### Existing multi-tissue RNA-seq dataset retrieval and processing

The multi-tissue dataset of human RNA-sequencing samples used to develop GeneNetwork was re-used and processed as described previously^6^. After pre-processing, this dataset contained 31,499 samples and 56,435 genes.

### Kidney-specific RNA-sequencing data retrieval

Meta-data of 3,108 publicly available kidney-derived RNA-sequenced samples was downloaded from the European Nucleotide Archive (ENA) on October 1, 2019. The keywords used to filter the data can be found in **supplementary table 1**. In addition, 86 samples from the Genotype-Tissue Expression (GTEx) Project were obtained using dbGaP accession number phs000424.v8.p2.

### Kidney-specific RNA-sequencing alignment

We used Kallisto^10^ to align the kidney expression dataset. The Kallisto index was based on Ensembl^11^ version 98 ncRNA and cDNA files (after removal of patch chromosomes) and made using default parameters, except for -k 31, and Kallisto version 0.43.1. Alignment was done using the Kallisto quant version 0.46.0 with default parameters and the addition of bootstrapping -b 30. For single-end data mapping, the additional parameters –l 200 and –s 20 were defined. The transcript counts were merged into gene counts. A TPM expression matrix was constructed based on the gene counts per sample.

### Kidney specific sample and gene selection

We excluded samples with ≤ 70% mapping reads, samples sequenced on platforms other than the Illumina sequencing platform and peripheral blood samples. The expression matrix with the remaining samples was quantile-normalized and log_2_-transformed, followed by principal component analysis (PCA)^12^ over the samples. Based on the first two principal components (PCs), a cut-off for good quality samples was set at 0.030 **(supplementary figure 1)**. We then excluded five of the samples that passed the PCA threshold but were sequenced using methods other than RNA-sequencing or ssRNA-sequencing, leaving 944 samples. Another 66 samples that correlated >0.9999 were removed because they were considered to be duplicates. The remaining 878 samples were used for further analysis **(supplementary figure 2)**.

The reads were aligned to 59,562 genes. We excluded 1,259 genes due to duplicate and/or no expression and 20 genes with zero variance, leaving 58,283 genes in the analysis.

### Sample clustering and investigation using UMAP

We investigated the remaining 878 RNA-sequencing samples using the UMAP clustering algorithm. UMAP values were generated using the umap() function in R version 3.5.1. We used PCs 1 and 2 as initial coordinates. Other parameters were defined as follows: n_threads = 24, n_epochs = 1000, n_neighbors = 100, min_dist = 0.1, init_sdev = 1e-4, learning_rate = 1, spread = 20, scale = “none” and nn_method = “fnn”. For all other parameters, default values were used. We plotted the UMAP values using base R plot function. Samples are colored based on literature descriptions of the studies included **(supplementary table 2)**.

### HPO alterations

For both the construction of KidneyNetwork and benchmarking and validation, we used gene− phenotype associations from HPO database^7^ version 1268. Annotation of genes to HPO-defined phenotypes is based on the gene−disease annotations in the OMIM^8^ morbid map (downloaded March 26, 2018) and the Orphanet^9^ “en_product6.xml” file version 1.3.1. Gene−disease annotations in these databases can be based on several factors, including statistical associations and large-scale copy number variations. To increase the accuracy of the gene−phenotype annotations used in KidneyNetwork, we filtered the gene−disease annotations for “The molecular basis for the disorder is known; a mutation has been found in the gene” for OMIM and for “Modifying germline mutation(s)”, “Disease-causing germline mutation(s)”, “Disease-causing somatic mutation(s)”, “Disease-causing germline mutation(s) (loss of function)”, or “Disease-causing germline mutation(s) (gain of function)” for Orphanet. We then rebuilt the gene−phenotype annotations in the HPO format that is used for network generation.

### Network generation

After sample and gene quality control (QC), the expression matrix of the remaining samples and genes was log_2_-transformed and gene counts were normalized using DESeq following the median of ratios method. Covariate analysis was done using Picard tools^13^. The covariate table was obtained using the command CollectRnaSeqMetrics with the following metrics: PCT_CODING_BASES, PCT_MRNA_BASES, PCT_INTRONIC_BASES, MEDIAN_3PRIME_BIAS, PCT_USABLE_BASES, INTERGENIC_BASES, INTRONIC_BASES, PCT_INTERGENIC_BASES and PCT_UTR_BASES. The option ‘strand_specificity’ was set to ‘first read transcription strand’ and the option ‘validation_stringency’ to ‘lenient’. The covariates were removed from the gene counts, and gene expression was correlated using Pearson correlation. The PC decomposition was performed over this correlation matrix.

### Decomposition

After filtering and QC of the entire dataset, the next step was to perform a decomposition to calculate the eigenvectors of the dataset. For this step, we used the PCA decomposition implemented by the Sklearn package^14^ version 0.22.2.post1 of the Python^15^ programming language version 3.6.3. The analysis was performed using the “full” svd_solver option. For both GeneNetwork and the gene regulatory network based on kidney-derived data, we defined the optimal number of components. Using explained variance cut-offs ranging between 0.3 and 0.8, the prediction accuracy (described in the next step) was calculated for pathways annotated in the HPO, Reactome^16^, KEGG^17^ and GO databases^18^ (Reactome, KEGG and GO databases downloaded on July 18, 2020). We chose to use the explained variance cut-offs that correspond to the highest average prediction accuracy, which were 0.5 for GeneNetwork and 0.7 for the network based only on kidney-derived data **(supplementary figures 3-4)**. Using these cut-offs, the first 165 eigenvectors for GeneNetwork and the first 170 eigenvectors for the kidney-derived data were identified and merged into a larger matrix containing all 335 eigenvectors. During this step, the 52,347 overlapping genes were kept.

### Gene−phenotype score calculation

The gene−phenotype score calculation was done in several steps **(supplementary figure 5)**. We first used the combined eigenvectors and the gene−phenotype annotations file as input. Phenotypes with fewer than 10 annotated genes in the HPO database were excluded. For each phenotype, a logistic model was fitted between the genes annotated for that phenotype against all genes annotated to at least one other phenotype in the annotation file. If genes of the current phenotype were also included in other phenotypes, we excluded these genes from the second gene set to fit the model. The model was fitted using the LogisticRegression class of the sklearn package with the ‘lbfgs’ solver, L2 regularization, a C-value of 1.0, a tolerance of 1e-6 and a max iteration of 6000. The model resulted in an intercept (β0) and β values corresponding to every component (β_1_ until β_336_). We used these β values and the eigenvector scores to calculate a gene log-odds score for every gene in the eigenvector table with the following formula:

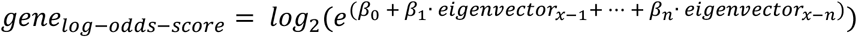

Where β_0_ is the intercept, β_1,…,_β_n_ are the β values of the logistic model and the eigenvector_x-1,..,_ eigenvector_x-n_ are the eigenvectors values of gene x.

To avoid overfitting of the gene log-odds-scores of already annotated genes, we fitted a new model for each annotated gene in which the gene−phenotype annotation was set to false for that gene. This means that the newly trained model does not incorporate the gene as a known gene for that phenotype. We used the corrected intercept and β values to calculate the gene log-odds-score for the annotated genes.

The log-odds were translated to gene z-scores by calculating a null distribution for each phenotype. We did this by random imputation of the gene labels of the eigenvector matrix and calculating the gene log-odds-score using the same formula and β values from the already-fitted models. These null-distributed gene scores were used to calculate the average and standard deviation of the gene log-odds-score. We subsequently calculated the gene z-scores with the formula:

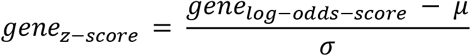

Where μ is the average and σ is the standard deviation of the gene log-odds-scores of the null distribution.

We calculated the null-distributed gene log-odds-scores of the genes annotated to a phenotype by using the intercept and β values obtained from the models calculated during the overfitting step.

To determine prediction accuracy, we calculated the area under the ROC-curve (AUC). The ROC-curve was calculated per phenotype using the predicted gene z-scores and known gene−phenotype annotations. The significance of the predictions was calculated using the two-sided Mann-Whitney rank test from the stats package of Scipy version 1.4.1. After Bonferroni-correction, a prediction was considered significant at p < 0.05.

### Comparing the prediction performances of KidneyNetwork and GeneNetwork

We compared the prediction performance of four distinct networks: (1) the original GeneNetwork, (2) the updated GeneNetwork, (3) the kidney-specific gene regulatory network based solely on kidney-derived samples and (4) the final KidneyNetwork combining the latter two. The quality of the HPO predictions made by these networks was assessed based on the AUC for each kidney-related phenotype, with kidney-related phenotypes defined as kidney-specific HPO terms **(supplementary table 3)**. Improved quality of a network was defined as improved prediction accuracy for kidney-related terms that were significantly predicted in each comparison of two networks and by an increased number of significantly predicted kidney-related terms. The significance of improvement in prediction accuracy of one network versus another was assessed using the DeLong test^19^ integrated in the pROC R package^20^.

### Applying KidneyNetwork to 13 kidney disease patients

One of the applications of KidneyNetwork is to prioritize candidate genes in patients with unsolved kidney disease. To evaluate this clinical application, we used KidneyNetwork to prioritize candidate genes for 13 patients with various kidney diseases using the GADO method^6^. GADO combines the gene prediction z-scores rendered through KidneyNetwork for a given set of HPO terms. When a given HPO term cannot be predicted significantly, GADO uses the parent term. Genes with a combined z-score ≥ 5 for the unique set of HPO terms associated with each patient were considered potential candidate genes for that patient.

The 13 patients included in the study are all suspected to have a monogenic kidney disease based on either family history, clinical presentation and/or early onset of their disease. However, no genetic cause had been found with diagnostic exome sequencing using trio-analyses or a diagnostic exome-based gene panel. Sequencing in these patients was performed at the University Medical Center Utrecht, as described previously^21^. All patients gave written informed consent for the use of their genetic data for research purposes. Based on their phenotype, HPO terms were assigned to these cases by two physicians from the genetics department. For each patient, the complete exome sequencing data were reanalyzed using CAPICE^22^ to identify potentially pathogenic variants. Genes containing variants with a minor allele frequency (MAF) < 0.005 and a recall ≥ 99%, corresponding with a mild CAPICE cut-off of ≥ 0.0027, were considered interesting candidates.

Overlapping the genes identified by the KidneyNetwork integration in GADO with those identified by CAPICE resulted in a list of genes for each patient. These genes and variants in these genes were manually reviewed by a nephrogenetics expert panel (AMvE, LRC, NVAMK) and differences in opinion were resolved through consensus discussion. Variants were assessed based on gnomAD allele frequency, the ClinVar database, the HGMD database, Grantham score and prediction tools (CADD, SIFT, MutationTaster, PolyPhen-2, Splice Prediction Module in Alamut Visual v.2.15). Genes containing potentially harmful variants were assessed based on available literature about that gene or related genes and on known gene−phenotype associations that could make this gene a more or less likely candidate. When autosomal recessive inheritance was expected based on the gene or the family history, and only one variant was found, the genetic data was reassessed for a second variant and SNP-array results were inspected for copy number variations. When sequencing data from parents was available, we assessed whether a variant was *de novo* or inherited. For the one resulting candidate gene, additional patients carrying variants in the same gene were identified via collaborators and the 100,000 Genomes Project. The GeneMatcher tool^23^ was used, and yielded no additional patients through February 17, 2021.

### 100,000 Genomes Project

Inclusion and genotyping of participants in the 100,000 Genomes Project, managed by Genomics England Limited (GEL), was previously described^24^. The multi-sample VCF dataset release v10, containing genome-wide sequencing data, was used to search for participants with a matching phenotype carrying rare variants in the candidate gene. We extracted high and moderate impact variants with a MAF < 0.001 and a scaled CADD-score (v1.5) of > 20 (or no CADD score if not applicable). For all participants who yielded a variant, we checked known kidney disease genes for (likely) pathogenic variants that might be causing their phenotype by checking whether a reportable variant was found in the diagnostic pipeline of the 100,000 Genomes Project and by checking for rare high/moderate impact variants in known renal genes defined by the complete diagnostic kidney gene panel of the University Medical Centre Utrecht NEF00v18.1 **(supplementary table 4)**.

## Results

### Data retrieval and sample clustering

3,108 publicly available kidney-derived RNA-sequencing samples and 86 kidney-derived GTEx RNA-sequencing samples were retrieved from the ENA database and the dbGaP database, respectively. After sample selection, 878 kidney-derived samples were used to develop KidneyNetwork **(supplementary figure 2)**. To investigate the remaining samples, we clustered and plotted them using the UMAP algorithm **(figure 1)**. Generally, the data clusters into three main clusters: primary non-tumor kidney data, kidney developmental samples and proximal tubule, glomerulus and renal cell carcinoma (RCC) samples. On the left side of the figure, clustering of pluripotent stem cell (PSC)-derived podocytes and PSC-derived organoids with primary fetal samples and nephron progenitor cells can be seen. On the right side, RCC samples cluster close to proximal tubule samples, and the RCC cluster closest to healthy primary tissue samples consists of non-clear cell RCC (nccRCC) samples. At the middle and bottom, healthy primary kidney samples cluster based on their tissue of origin.

**Figure 1:**
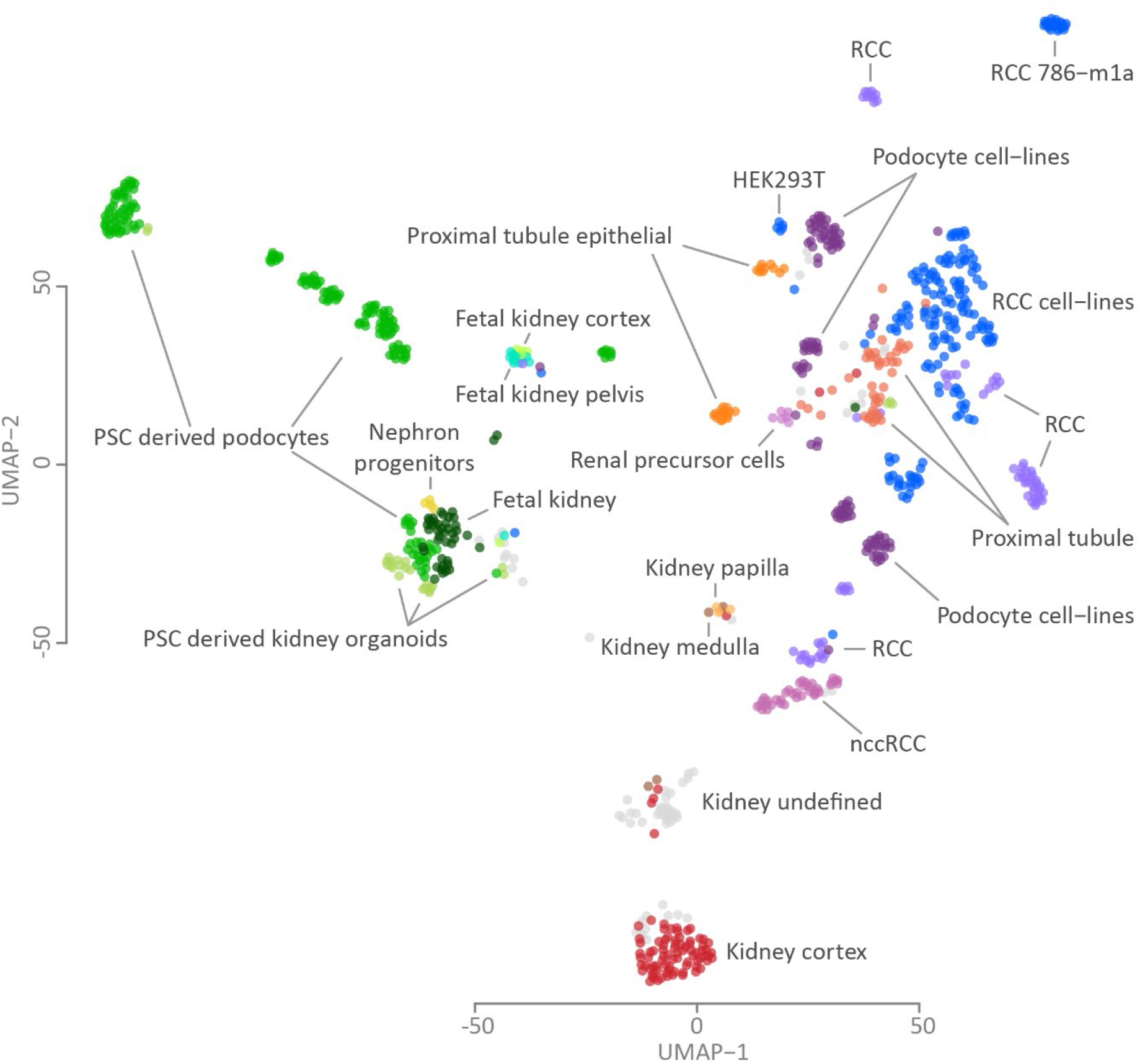
UMAP visualization of the kidney-derived expression data. 878 samples group into three main clusters: healthy primary tissue (middle and bottom), developmental samples (left) and renal cell carcinoma (RCC) samples (right).

### KidneyNetwork improves gene−phenotype predictions

We first updated GeneNetwork with the updated HPO database **(supplementary figure 6)** and optimized the gene network building pipeline **(supplementary figure 7)**. These changes yielded an improvement in the general GeneNetwork compared to the previous version **(supplementary figure 8)**. We then used the improved pipeline to build the kidney-specific gene regulatory network. As expected given the small sample size, this version of the kidney-specific network performed less well than GeneNetwork **(supplementary figure 9)**. Subsequently combining GeneNetwork and the kidney specific gene co-expression network into KidneyNetwork yielded our best results for kidney-related HPO terms **(figure 2A; supplementary table 5)**.

**Figure 2:**
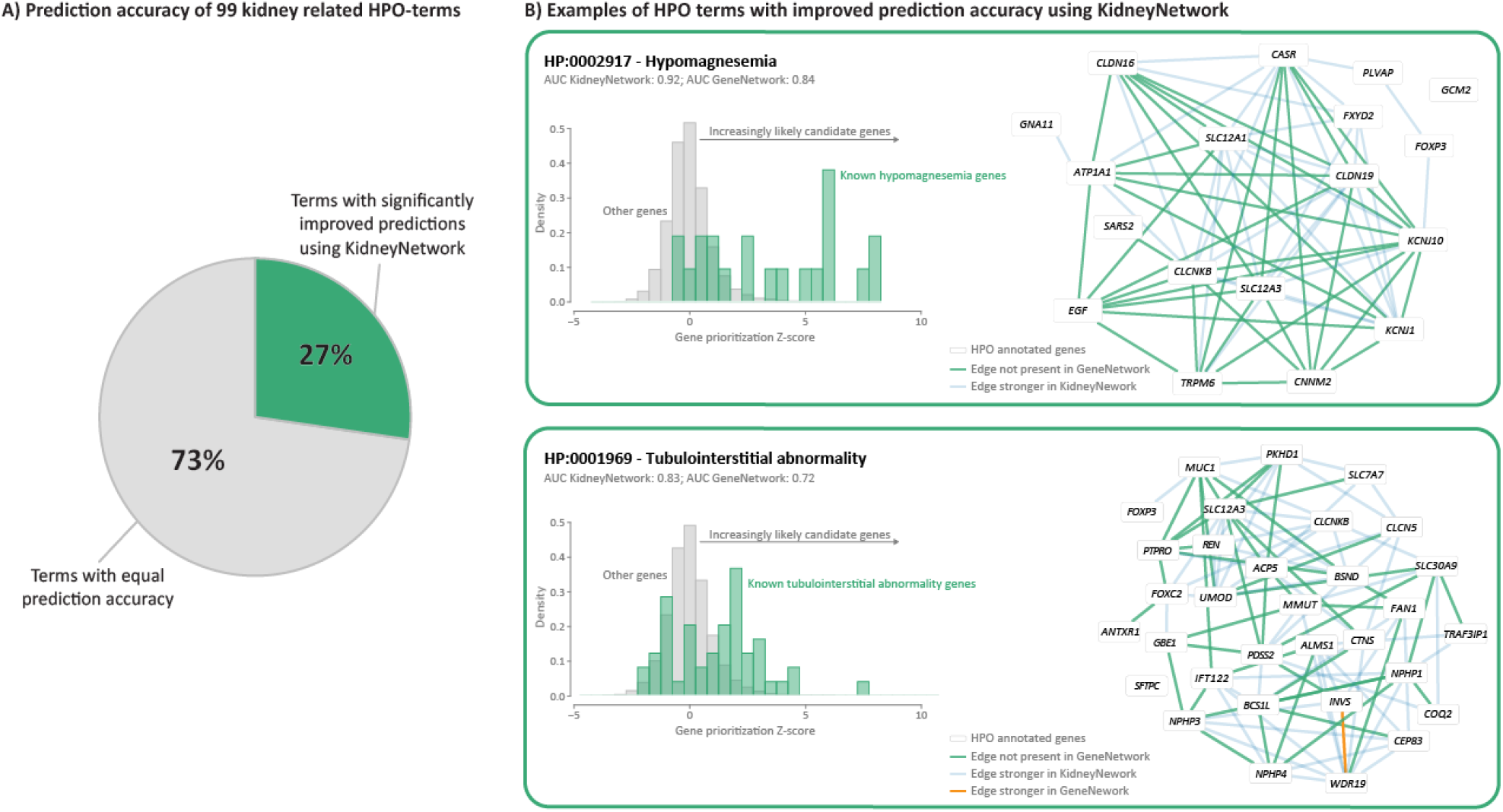
KidneyNetwork performs better for kidney-related HPO terms than the updated GeneNetwork. A) 27% of kidney-related phenotypes are predicted significantly better using KidneyNetwork, as compared to GeneNetwork. B) Density plots of the gene prediction scores within two of the most improved phenotypes, hypomagnesemia and tubulointerstitial abnormality, show higher prediction values for the genes annotated for the phenotype and also predict potential unknown candidate genes. The networks predicted using KidneyNetwork shows more and stronger correlations between the annotated genes than the networks predicted using GeneNetwork.

We calculated the number of pathways with a significant improvement in prediction accuracy for KidneyNetwork compared to GeneNetwork using the DeLong test^19^. For this analysis, phenotypes were grouped into kidney-related phenotypes and non-kidney-related phenotypes. Within the kidney-related phenotypes, no phenotypes were significantly better predicted in GeneNetwork compared to KidneyNetwork. In contrast, 27% of kidney-related pathways were significantly better predicted by KidneyNetwork compared to GeneNetwork **(figure 2A)**.

Two examples of improved kidney-related HPO terms are hypomagnesemia and tubulointerstitial abnormality **(figure 2B)**. Visualization of these phenotypes in density plots shows higher prioritization z-scores for known disease-related genes compared to non-annotated genes. For unknown genes, the higher the prediction z-score, the more likely it is to be a candidate disease gene. Visualizing the gene interaction networks of known disease genes based on the prediction scores again shows the increase in the number and strength of interactions obtained using KidneyNetwork compared to GeneNetwork.

To compare the performances of GeneNetwork and KidneyNetwork, we performed a paired t-test over the 27% significantly improved kidney-related HPO terms and found an overall significant improvement in performance of KidneyNetwork (mean AUC: 0.83) compared to GeneNetwork (mean AUC: 0.78) (t-test p-value: 1.2 × 10^−8^). This indicates that, overall, kidney-related terms can be predicted with a higher accuracy using KidneyNetwork compared to GeneNetwork.

We also saw an increase in the number of significant predicted kidney-related HPO terms for KidneyNetwork (n=71) compared to GeneNetwork (n=63). This led us to hypothesize that KidneyNetwork predicts kidney-related terms with higher accuracy overall and is therefore capable of predicting more kidney-related phenotypes with higher significance. To test this, we conducted a paired t-test considering all kidney-related HPO terms. Overall, there was a significant difference between the HPO AUC scores in GeneNetwork (mean AUC: 0.74) and KidneyNetwork (mean AUC: 0.76) (t-test p-value: 4.5 × 10^−8^). This result suggests that KidneyNetwork predicts more kidney-specific HPO terms with a higher prediction accuracy than GeneNetwork.

### KidneyNetwork predicts *ALG6* as a candidate gene for kidney cysts and liver cysts

To examine the clinical utility of KidneyNetwork, we applied GADO^6^ to data from 13 patients with a suspected hereditary kidney disease but no genetic diagnosis, using KidneyNetwork as the input matrix. For each patient, we identified which genes prioritized by GADO with KidneyNetwork overlap with genes containing potentially pathogenic variants predicted by CAPICE^22^. The resulting gene lists contained 1−4 candidate genes for 9 of the 13 patients **(supplementary table 6)**. In one patient (SAMPLE6), manual curation of this list identified *ALG6* (ALG6 alpha-1,3-glucosyltransferase) as a potential candidate gene to explain the patient’s renal and hepatic cysts **(figure 3)**. The combined z-score for *ALG6* for the imputed HPO terms was significant in KidneyNetwork after multiple testing correction (z = 5.43). This gene would have been missed when we had used the updated GeneNetwork: there *ALG6* did not reach the significance threshold of z-score ≥ 5.

**Figure 3:**
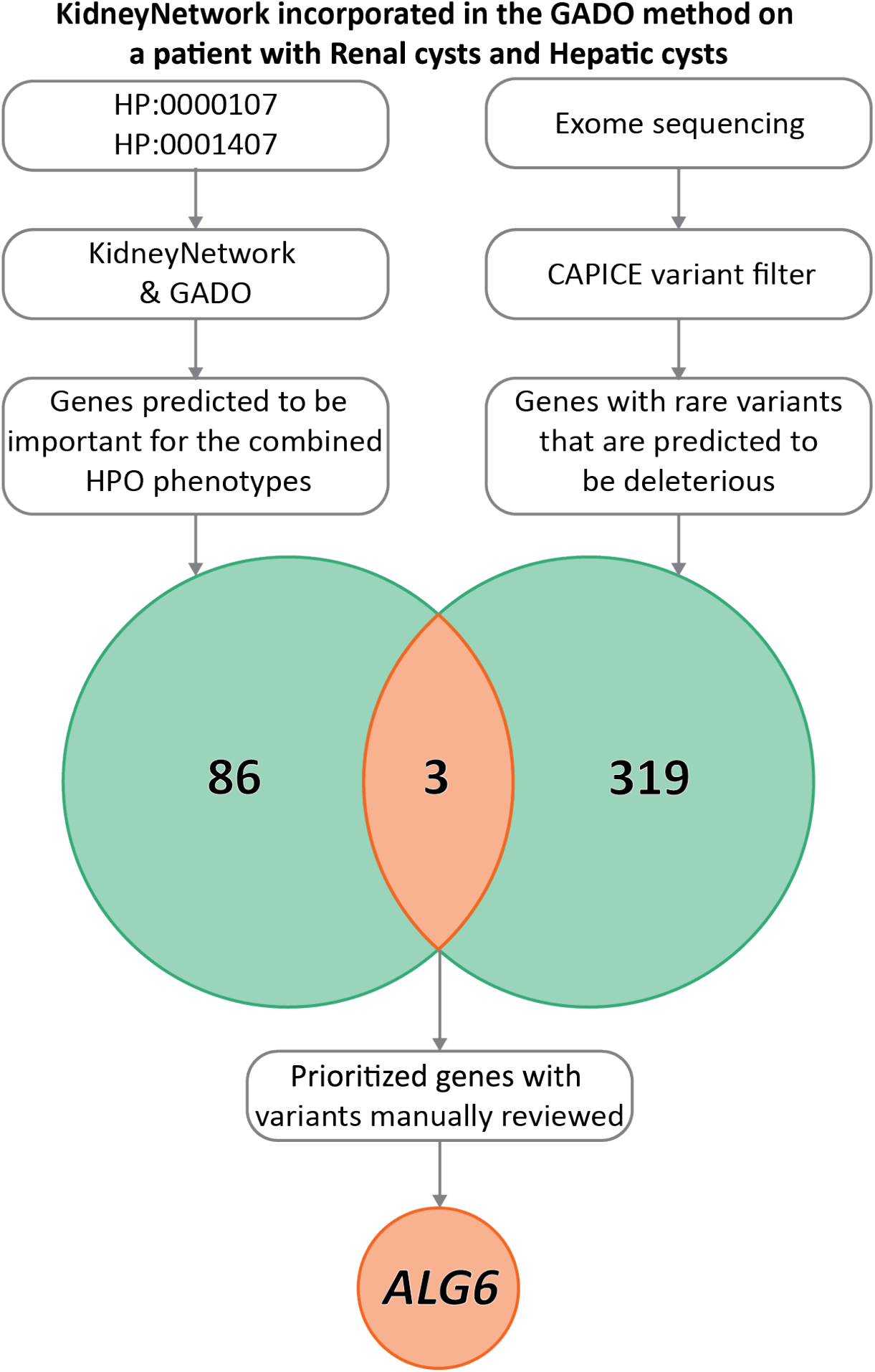
KidneyNetwork incorporated in the GADO method in a patient with renal and hepatic cysts. 89 candidate genes out of all genes were prioritized by KidneyNetwork using GADO, based on the HPO terms “Renal cysts” (HP:0000107) and “Hepatic cysts” (HP:0001407). Exome-sequencing data interpretation method CAPICE yielded 322 genes containing potentially pathogenic variants in the patient’s exome sequencing data. When overlapping these gene lists three genes were identified that met the selection criteria, one being ALG6.

### *ALG6* as candidate gene for patients with renal and hepatic cysts

The *ALG6* variant c.680+2T>G carried by SAMPLE6 is heterozygous. This is a known pathogenic splice site variant that results in congenital disorder of glycosylation (CDG) type Ic when pathogenic variants are present on both alleles^25,26^. *ALG6* strongly resembles *ALG8* which has been implicated in kidney and liver cyst phenotypes^27^, and according to KidneyNetwork, *ALG6* and *ALG8* are highly co-regulated (z-score = 8.59). We believed this variant could be causative for this patient’s phenotype and searched for additional patients with *ALG6* variants who displayed similar phenotypes.

Through the 100,000 Genomes Project dataset and collaborators, we identified three additional patients with kidney and/or liver cysts carrying a heterozygous potentially deleterious variant in *ALG6* that could be causative for their phenotype. **Table 1** lists the phenotypes and detected variants. Including SAMPLE6, we identified three patients with known splice site variants that were reported to be disease-causing in severely affected CDG patients upon compound-heterozygosity. In contrast, our patients presented with a mild phenotype of multiple kidney cysts and/or liver cysts (**figure 4**), but no eGFR decline was reported despite advanced age. All patients are currently between 48-70 years old. The only patient with a novel missense variant displayed a more severe phenotype with progression to kidney failure. No (likely) pathogenic variant was found in any known renal disease genes in these patients. Additional patients with rare *ALG6* variants deemed non-causative for various reasons are listed in **supplementary table 7**.

**Table 1:** Clinical information on patients with heterozygous ALG6 variants, including variant details and in silico predictions.

| patient (gender) | phenotype | family history | variant nomen (cDNA)<sup>3</sup> | variant nomen (protein) | zygosity | allele frequency gnomAD v2.1.1 | CADD score (PHRED) v1.5 | reference for variant |
| --- | --- | --- | --- | --- | --- | --- | --- | --- |
| SAMPLE6 (female) | multiple renal cysts and multiple hepatic cysts (incidental finding), normal eGFR | child and sibling with renal cysts<sup>1</sup> | c.680+2T>G | splice site variant | heterozygous | 8.02e-6 | 29.3 | Morava et al, Sun et al<sup>25,26</sup> |
| GEL1 (male) | multiple renal cysts, gout, CKD stage 5 at age 53 | no affected family members<sup>2</sup> | c.68T>A | p.Leu23His | heterozygous | 0 | 26.1* | novel |
| GEL2 (male) | multiple renal cysts | no affected family members<sup>2</sup> | c.257+2dup | splice site variant | heterozygous | 1.59e-5 | 25.2 | Newell et al<sup>28</sup> |
| LE1 (female) | mild polycystic liver disease without renal manifestation, normal eGFR | sibling with liver cysts<sup>1</sup> | c.257+5G>A | splice site variant<sup>#</sup> | heterozygous | 4.72e-4 | 22.0 | Imbach et al, Westphal et al, Drijvers et al<sup>29-31</sup> |

<sup>1</sup>Segregation pending; <sup>2</sup>No DNA available for segregation; <sup>3</sup>NM\_013339.3; \*additional prediction scores (SIFT, PolyPhen-2, MutationTaster, Grantham Distance) are listed in supplementary table 8, <sup>#</sup>splice site analysis presented in supplementary figure 10, | CKD=chronic kidney disease, NA=not applicable

**Figure 4:**
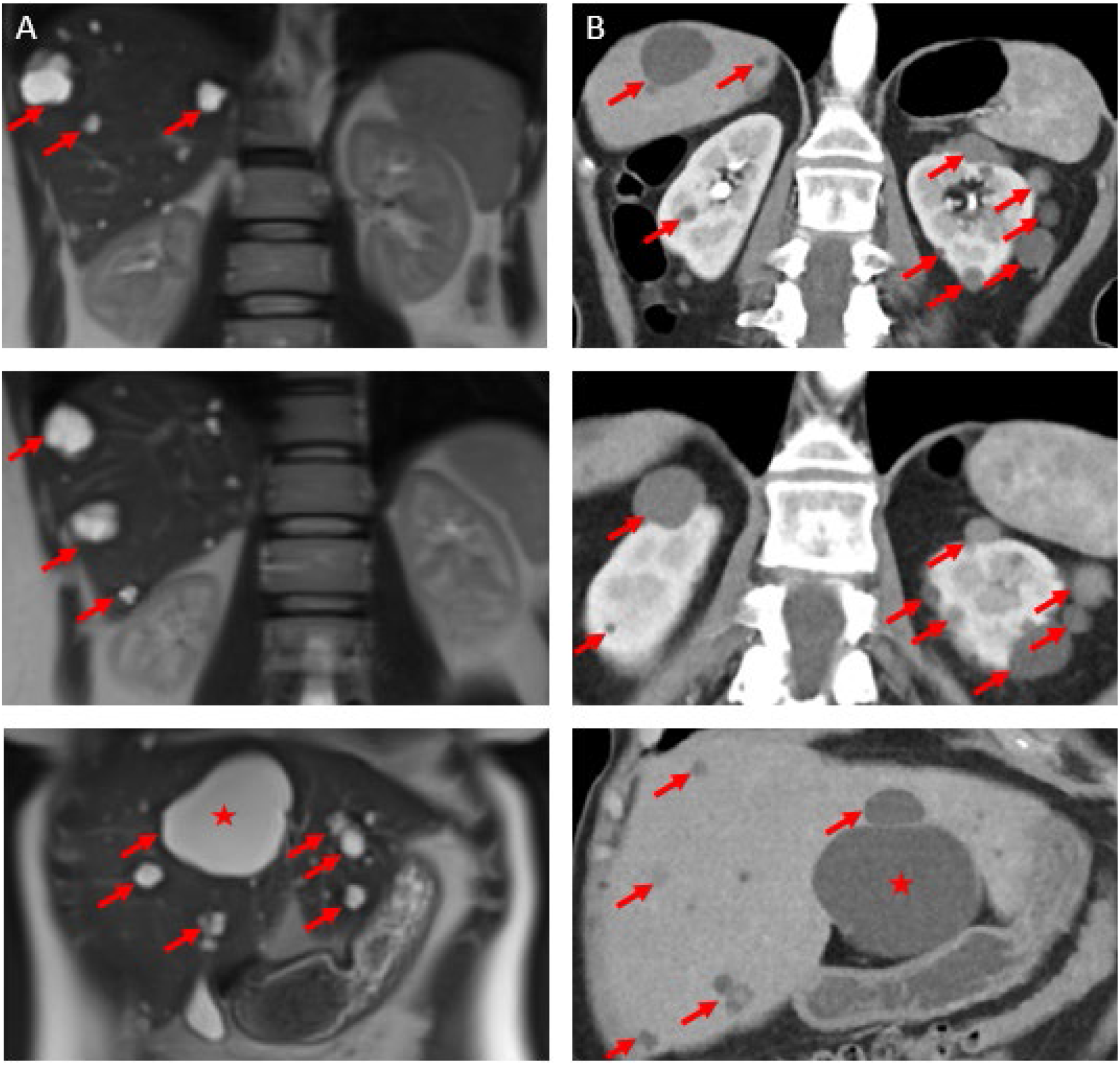
Imaging from two patients. A) abdominal MRI illustrating polycystic liver disease in LE1. Hepatic cysts are highlighted by red arrows, with the largest cyst located in liver segment IV (red asterisk), necessitating surgical intervention for progressive cholestasis. Of note, both kidneys presented with normal morphology in absence of any cystic lesions. B) abdominal CT illustrating polycystic kidneys and liver in SAMPLE6. Cysts are highlighted by red arrow, with the largest hepatic cyst measuring 7.7 cm (red asterisk).

## Discussion

Although advances in exome sequencing can aid in the genetic diagnosis of kidney disease patients, a significant proportion of patients with a suspected genetic cause remain without a genetic diagnosis. When diagnostic exome or genome sequencing is performed, interpretation is often limited to known kidney disease genes. Even with exome-wide analysis, many patients remain without a genetic diagnosis because the function of a large proportion of genes remains unclear. Therefore, identifying which genes are involved in kidney disease is essential for improving the diagnostic yield in kidney disease patients. To improve this situation, we developed KidneyNetwork, a kidney-specific co-expression network that can be used to prioritize previously unknown kidney disease genes.

KidneyNetwork combines co-expression with information on known gene−phenotype annotation to predict which previously unknown genes may also play a role in phenotype etiology. KidneyNetwork was built using two human RNA-sequencing datasets: a kidney sample dataset and the previously published multi-tissue dataset used to build GeneNetwork. Combining these datasets into KidneyNetwork improved phenotype predictions related to kidney disease when compared to networks based on the datasets separately. As proof of principle, we used KidneyNetwork to prioritize variants from exome sequencing in a small group of suspected hereditary kidney disease patients with diverse phenotypes, and this identified *ALG6* as a candidate gene for kidney and liver cysts.

We conclude that *ALG6* is a credible candidate gene for kidney and liver cysts because of its resemblance to known disease genes in the same pathway. *ALG6* and *ALG8* are both members of the α3-glucosyltransferase family^32^. In addition to *ALG8*^27^, *ALG9* heterozygous variants have recently also been implicated in the etiology of kidney and liver cyst phenotypes^33^. These three genes are closely related in a biosynthetic pathway for lipid-linked oligosaccharides^34^. Interestingly, recessive loss-of-function variants in both *ALG8* and *ALG9* result in CDG, with kidney and/or liver cysts described. Cysts have not (yet) been described for *ALG6-*CDG. The classic *ALG6*-CDG phenotype is characterized by developmental delay, failure to thrive and multiple neurological symptoms such as hypotonia, ataxia, proximal muscle weakness and epilepsy^26^. The phenotype present in SAMPLE6, GEL2 and LE1 is similar to the relatively mild phenotype described in patients carrying a heterozygous *ALG8* or *ALG9* pathogenic variant. *ALG6* has previously been suggested to be involved in one individual with autosomal dominant polycystic kidney disease (ADPKD)^35^. However, that patient, who carried two missense variants with inconclusive predictions that have not been functionally assessed, had a very severe phenotype that did not match the expected phenotype for *ALG6*. In GEL1, the only patient in our analysis carrying a missense variant, we also see a relatively severe phenotype with kidney failure needing replacement therapy at age 53. This predicted deleterious missense variant is not found in gnomAD, and the patient’s severe phenotype might be explained by variability in phenotype or a specific effect of this variant (e.g. gain-of-function). Variability in phenotype severity has been observed in patients carrying heterozygous *ALG9* variants, but only one patient has been described with kidney failure and no conclusions could be drawn in this case due to lack of records and imaging before kidney failure commenced^36^. Another explanation for GEL1’s phenotype is that we might have missed another deleterious variant despite exome-wide analysis. Functional experiments are thus needed to unravel the exact cellular function of ALG6 in kidney cyst pathogenesis and to prove the pathogenic effect of all these variants.

### Strengths and limitations

We realize that *ALG6* based on the present literature alone would be a candidate gene for the cyst phenotype in SAMPLE6. However, this only proves the strength of our method; out of 322 genes with potentially deleterious variants this true candidate gene was prioritized to the top 3, making going into exome-wide sequencing data − for more patients, with various phenotypes − time-efficient and worthwhile.

Building gene co-expression networks requires a large number of RNA-sequencing samples^6^ in order to achieve accurate function predictions, numbers that are not often available for specific tissues. To overcome this issue, one earlier approach used hierarchical similarities between tissue types^37^. However, this solution requires a priori gene selection due to its computational burden. In contrast, our method can be used to make unbiased genome-wide predictions. Moreover, the hierarchical approach would have to be repeated for each new tissue of interest, whereas the multi-tissue dataset can be re-used to build a different tissue-specific network using our method. Another earlier approach used differential expression between different tissue types^38^. In this approach, the top 10% most differentially expressed genes were correlated with kidney-related GWAS loci. Using differential expression allows predictions to be made regardless of previous knowledge on gene− phenotype interactions. However, this also requires applying a differential expression cut-off, which was set to the top 10% most differentially expressed genes, for inclusion. In contrast, our approach makes use of underlying biological structures in RNA-sequencing data to obtain a prediction score for every gene. While combining differential expression with GWAS summary statistics allows for unbiased gene predictions, the reliability of experimentally validated HPO annotations is higher than that of GWAS results, and integrating the HPO database thus results in more reliable predictions. Moreover, we make simultaneous predictions for all HPO terms, whereas the GWAS-based approach needs to be repeated for each GWAS of interest.

Combining kidney-specific RNA-sequencing samples with the multi-tissue dataset allowed us to overcome both the issue of sample size and the challenges in observing tissue-specific differential expression when using only tissue-specific expression datasets. In addition, during the development of KidneyNetwork we did not have to limit the number of genes that the network is built upon. Furthermore, KidneyNetwork users can get predictions for all possible genes in an unbiased approach, and gene prioritizations for a combination of HPO terms can be obtained.

A downside of using RNA-sequencing data from bulk samples is that we have limited power to make inferences for lowly expressed genes, which is particularly important for genes that are specific to rare cell-types. However, as more cell-type specific and single-cell RNA-sequencing data becomes available in the future, creating co-expression networks based on different kidney cell types might solve this for genes that are expressed more abundantly within specific cell types. Another limitation of using only RNA-sequencing data is that other biological processes potentially involved in disease development, for example post-translational modifications and protein-protein interactions, are currently not considered by our prediction model.

### Improved gene function predictions

We show that our improved method for assigning gene functions and HPO terms to genes outperforms our previously published model. Our leave-one-out cross validation approach ensures that predictions are not overfitted, that the reported AUC values are not inflated and that our method is robust.

Furthermore, before predicting gene−phenotypes associations, we excluded gene−disease associations from the HPO database that had little experimental evidence, because prediction accuracy is dependent on the accuracy of annotated gene−phenotype associations. Prediction accuracy is based on the true positive gene predictions and true negative gene predictions, which means that the more accurately the known genes are mapped to phenotypes, the better the predictions will be. We noticed that not all genes mapped to HPO terms were mapped based on experimental validation, with some of the associations based on other factors such as statistical associations. As these associations could introduce noise into the network predictions, they were not considered true positives in our analysis. Another observation was made for phenotypes belonging to diseases with large duplications or deletions: Genes positioned in the deleted or duplicated areas are currently annotated to these diseases in the HPO database and thus linked to the disease associated−phenotypes. As these genes not necessarily cause such phenotypes, our network predictions would have been affected by these genes. We therefore only included gene− phenotype associations based on known associations in the analysis. Gene-phenotype association accuracy will improve once more genes are annotated and validated for each phenotype. Therefore, we expect an improvement in network prediction accuracy as gene−phenotype association knowledge increases and is added to the HPO database, and we expect this will help to establish more genetic diagnoses for kidney disease patients in the future.

### Applications of KidneyNetwork

We have developed https://kidney.genenetwork.nl/ through which we provide the gene-HPO term prediction. Using the same prediction algorithm that we used to assign genes to HPO-terms, we also predicted which genes are likely to be involved in GO, KEGG and Reactome pathways.

Here we also provide an online version of GADO that can be used to prioritize relevant genes for patients with a suspected rare kidney disease. It is possible to specify the phenotype of a patient using HPO terms and provide a list of genes harboring potential disease-causing variants. These genes will then be ranked using KidneyNetwork, thereby allowing the identification of which genes are more likely to be involved in the patient’s disease. Since it is not necessary to upload personal genetic information, this method respects patient privacy.

### Future directions

Application of KidneyNetwork to unsolved cases from diagnostics, large research cohorts and, for instance, GWAS datasets will result in more insight into kidney physiology and pathophysiology. To further improve the accuracy of kidney phenotype prediction, we plan to incorporate single-cell RNA-sequencing data, which we expect will yield more detailed and accurate gene−phenotype predictions.

## Conclusion

We present KidneyNetwork, a co-expression network that can help increase the genetic diagnostic yield in kidney disease patients. The method we developed to combine multi-tissue data with tissue-specific data can easily be extended to other tissues, allowing improved predictions for other tissue-specific diseases. Using KidneyNetwork, we identified *ALG6* as candidate gene for the phenotypes renal and/or hepatic cysts and therefore add *ALG6* as a credible candidate for the ADPKD/PCLD spectrum. KidneyNetwork provides a useful tool to help with the interpretation of genetic variants. It can therefore be of great value in translational nephrogenetics and ultimately improve the diagnostic yield in kidney disease patients.

## Data Availability

To make the gene-co-regulation-based HPO predictions publicly available a website was constructed: https://kidney.genenetwork.nl/. Informed consent did not cover uploading exome sequencing data from patients.

## Author contributions

P.D., L.F., A.M.v.E, F.B. and L.R.C. designed the study. N.V.A.M.K provided structural feedback on the study design and progress. F.B., F.S. and H.W. built KidneyNetwork. N.d.K. and H-J.W. provided help and feedback with the process of building KidneyNetwork. F.B. and H.W. did the benchmarking analysis. S.L. did the Capice analysis. F.B. and P.D. did the GADO analysis. A.M.v.E., N.V.A.M.K. and L.R.C. manually assessed results from GADO analysis. L.R.C. did the literature study for *ALG6* and the genomic analysis. L.R.C., J.H., D.S., R.S., A.M.v.E. and B.v.d.Z. analysed the *ALG6* variants. R.O. built and maintained the KidneyNetwork website. H.W., P.D. and F.B. made the figures. P.D., L.F. A.M.v.E., B.v.d.Z., J.H., F.v.R., R.O., F.S., N.d.K., H-J.W. and S.L. critically assessed the paper. L.R.C., F.B. and H.W. drafted and revised the paper. All authors approved the final version of the manuscript.

## Acknowledgements

We thank the study participants and their families for their contributions. We also thank the UMCG Genomics Coordination center, the UG Center for Information Technology, and their sponsors BBMRI-NL & TarGet for storage and computing infrastructure. We specifically thank Sido Haakma and Erik Schaberg for providing and setting up the virtual machine on which the KidneyNetwork website is hosted. We also thank Robert Ernst, Hanneke van Deutekom and Gijs van Haaften for helpful discussions and technical support and Katherine McIntyre for editing the manuscript. This work was supported by the Dutch Kidney Foundation (18OKG19). Several authors of this publication are members of the European Reference Network for Rare Kidney Diseases (ERKNet) Project ID No 739532. This research was made possible through access to the data and findings generated by the 100,000 Genomes Project. The 100,000 Genomes Project is managed by Genomics England Limited (a wholly owned company of the Department of Health and Social Care). The 100,000 Genomes Project is funded by the National Institute for Health Research and NHS England. The Wellcome Trust, Cancer Research UK and the Medical Research Council (MRC) have also funded research infrastructure. The 100,000 Genomes Project uses data provided by patients and collected by the National Health Service as part of their care and support.

The Genotype-Tissue Expression (GTEx) Project was supported by the Common Fund of the Office of the Director of the National Institutes of Health (commonfund.nih.gov/GTEx). Additional funds were provided by the NCI, NHGRI, NHLBI, NIDA, NIMH, and NINDS. Donors were enrolled at Biospecimen Source Sites funded by NCI\Leidos Biomedical Research, Inc. subcontracts to the National Disease Research Interchange (10XS170), Roswell Park Cancer Institute (10XS171), and Science Care, Inc. (X10S172). The Laboratory, Data Analysis, and Coordinating Center (LDACC) was funded through a contract (HHSN268201000029C) to the The Broad Institute, Inc. Biorepository operations were funded through a Leidos Biomedical Research, Inc. subcontract to Van Andel Research Institute (10ST1035). Additional data repository and project management were provided by Leidos Biomedical Research, Inc.(HHSN261200800001E). The Brain Bank was supported supplements to University of Miami grant DA006227. Statistical Methods development grants were made to the University of Geneva (MH090941 & MH101814), the University of Chicago (MH090951,MH090937, MH101825, & MH101820), the University of North Carolina -Chapel Hill (MH090936), North Carolina State University (MH101819),Harvard University (MH090948), Stanford University (MH101782), Washington University (MH101810), and to the University of Pennsylvania (MH101822). The datasets used for the analyses described in this manuscript were obtained from dbGaP at http://www.ncbi.nlm.nih.gov/gap through dbGaP accession number phs000424.v8.p2.

## Supplemental material

### Table of Contents

#### Supplemental Figures

Supplementary figure 1. Principal component analysis (PCA) was used for sample quality control and selection

Supplementary figure 2. Flowchart visualizing the sample selection of kidney-derived RNA sequencing data

Supplementary figure 3: Selecting eigenvector cut-off value for GeneNetwork data based on highest average AUC value of different database

Supplementary figure 4: Selecting eigenvector cut-off value for kidney-derived samples data based on highest average AUC of different databases

Supplementary figure 5. Pipeline of the updated network analysis

Supplementary figure 6. AUC value comparison between the original GeneNetwork and GeneNetwork based on the updated HPO database

Supplementary figure 7. AUC value comparison between the original GeneNetwork pipeline and the updated GeneNetwork pipeline with the original HPO data

Supplementary figure 8. AUC value comparison between the original GeneNetwork and the updated GeneNetwork

Supplementary figure 9: AUC value comparison of kidney-related HPO terms between GeneNetwork and the network created with kidney-derived data

Supplementary figure 10. *ALG6*-splice site analysis c.257+5G>A

#### Supplemental Tables

Supplementary table 1. Keywords for kidney sample selection

Supplementary table 2. Sample annotations

Supplementary table 3. Kidney-specific HPO terms

Supplementary table 4. Diagnostic kidney gene panel (n=379) of the University Medical Centre Utrecht NEF00v18.1

Supplementary table 5. Improved kidney-specific HPO terms

Supplementary table 6. Phenotypes and genes in 13 kidney disease patients

Supplementary table 7. Information on patients with *ALG6* variants deemed non-causative, including variant details and in silico predictions

Supplementary table 8. Additional prediction scores for *ALG6* variant in patient GEL1

