## Supplementary data for "KidneyNetwork: Using kidney-derived gene expression data to predict and prioritize novel genes involved in kidney disease"

### Supplemental material

**Supplementary figure 1: Principal component analysis (PCA) was used for sample quality control and selection.** A cut-off of 0.030 was chosen, and all samples above the threshold were considered good quality. Right figure is based on the same data as the left figure but has an expanded x-axis to produce a more detailed plot.

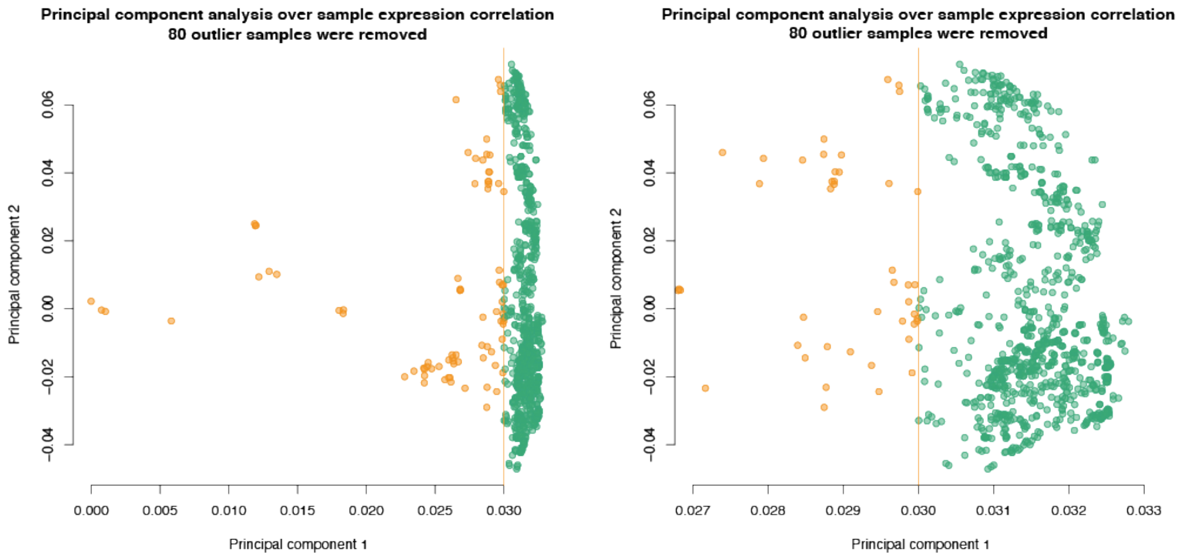

**Supplementary figure 2: Flowchart visualizing the sample selection of kidney-derived RNA sequencing data.** RNA-seq = RNA-sequencing, ENA = European Nucleotide Archive, dbGaP = database of Genotypes and Phenotypes, PCA = principal component analysis, ssRNA-seq = strand-specific RNA sequencing

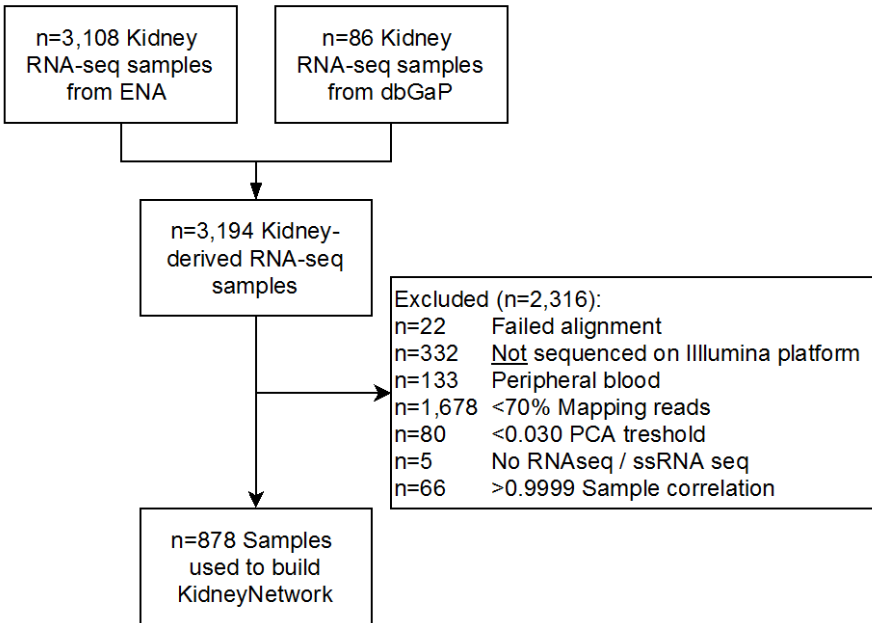

**Supplementary figure 3: Selecting eigenvector cut-off value for GeneNetwork data based on highest average AUC value of different database.** An explained variance cut-off of 0.5 gives the highest prediction accuracy for GeneNetwork, averaged over several databases. Each colored line represents the mean AUC of the prediction of all pathways within one database. Black line represents the average AUC over all the databases included. The number of eigenvectors included was based on the explained variance cut-off chosen.

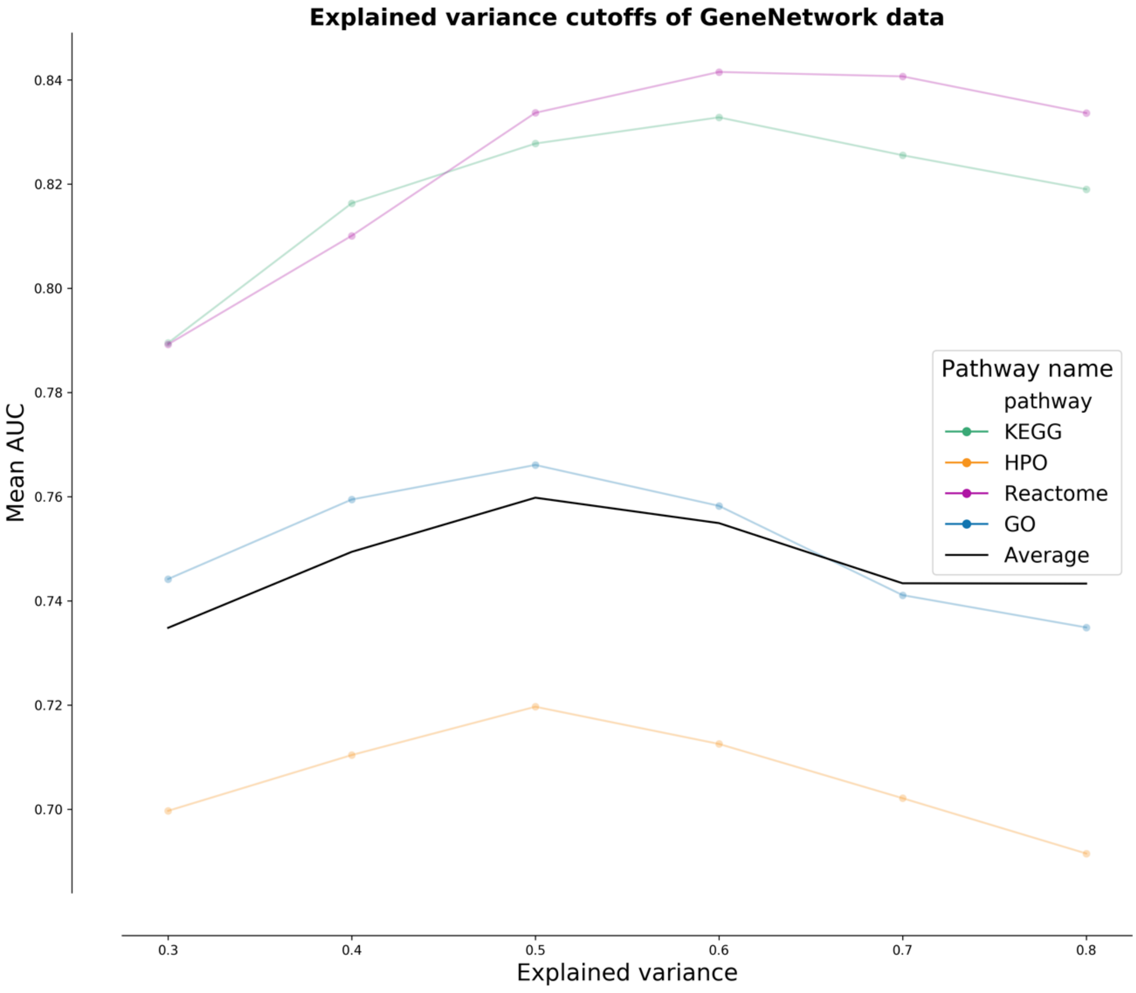

**Supplementary figure 4: Selecting eigenvector cut-off value for kidney-derived samples data based on highest average AUC of different databases.** An explained variance cut-off of 0.7 gives the highest prediction accuracy for the kidney-specific gene regulatory network based solely on 898 kidney-derived samples, averaged over several databases. Colored lines represent the mean AUC of the prediction of all pathways within one database. The black line represents the average AUC over all included databases. The number of eigenvectors included was based on the explained variance cut-off chosen.

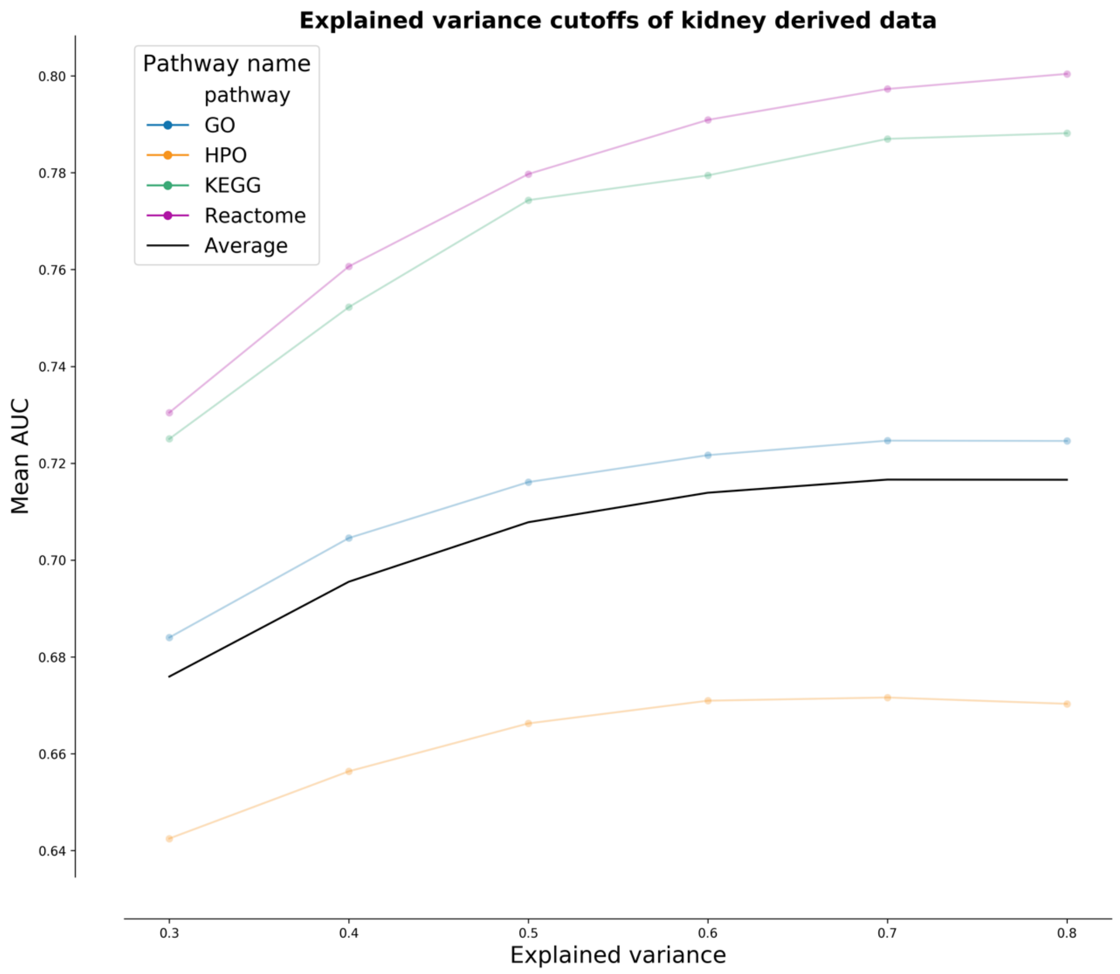

**Supplementary figure 5: Pipeline of the updated network analysis.** We start with a decomposition step of the input dataset to identify the components. These components are then used to fit a logistic model for each phenotype where the intercept and coefficients are used to calculate a gene log-odds score using the input components. The gene log-odds scores are translated to z-scores by permutation of the gene-components matrix to create a null distribution. This null distribution is used to calculate the gene log-odds scores of the null distribution, and the mean and standard deviation of these values are used to calculate the z-score for each gene.

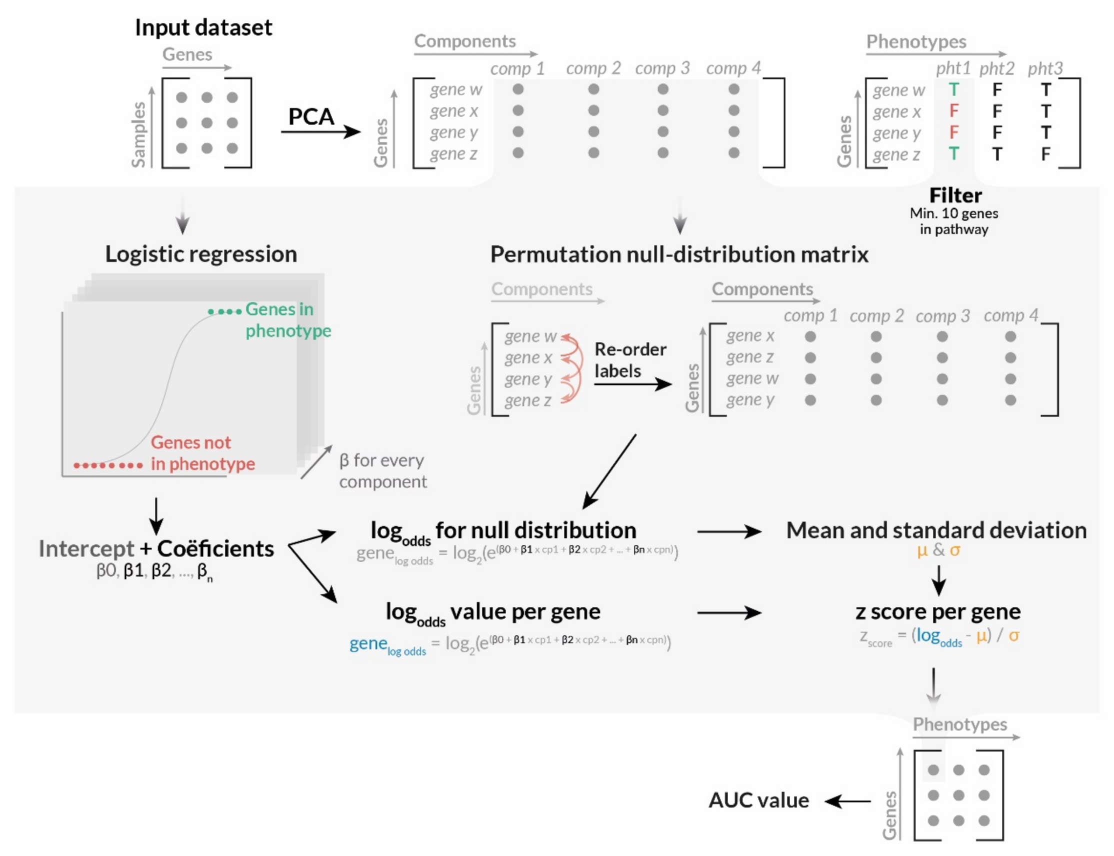

**Supplementary figure 6: AUC value comparison between the original GeneNetwork and GeneNetwork based on the updated HPO database.** The prediction accuracy of the original GeneNetwork pipeline described in the GADO paper (x-axis) compared to the same pipeline with the updated HPO database described in this paper (y-axis). Every dot represents the AUC value of one HPO term. Color scale is based on the significance of the prediction in one, both, or neither method after multiple testing correction. The mean AUC of the original GeneNetwork is 0.69, and 1,751 pathways are predicted to be Bonferroni significant. The mean AUC of GeneNetwork with updated HPO data is 0.70, and 2,059 pathways are predicted to be Bonferroni significant.

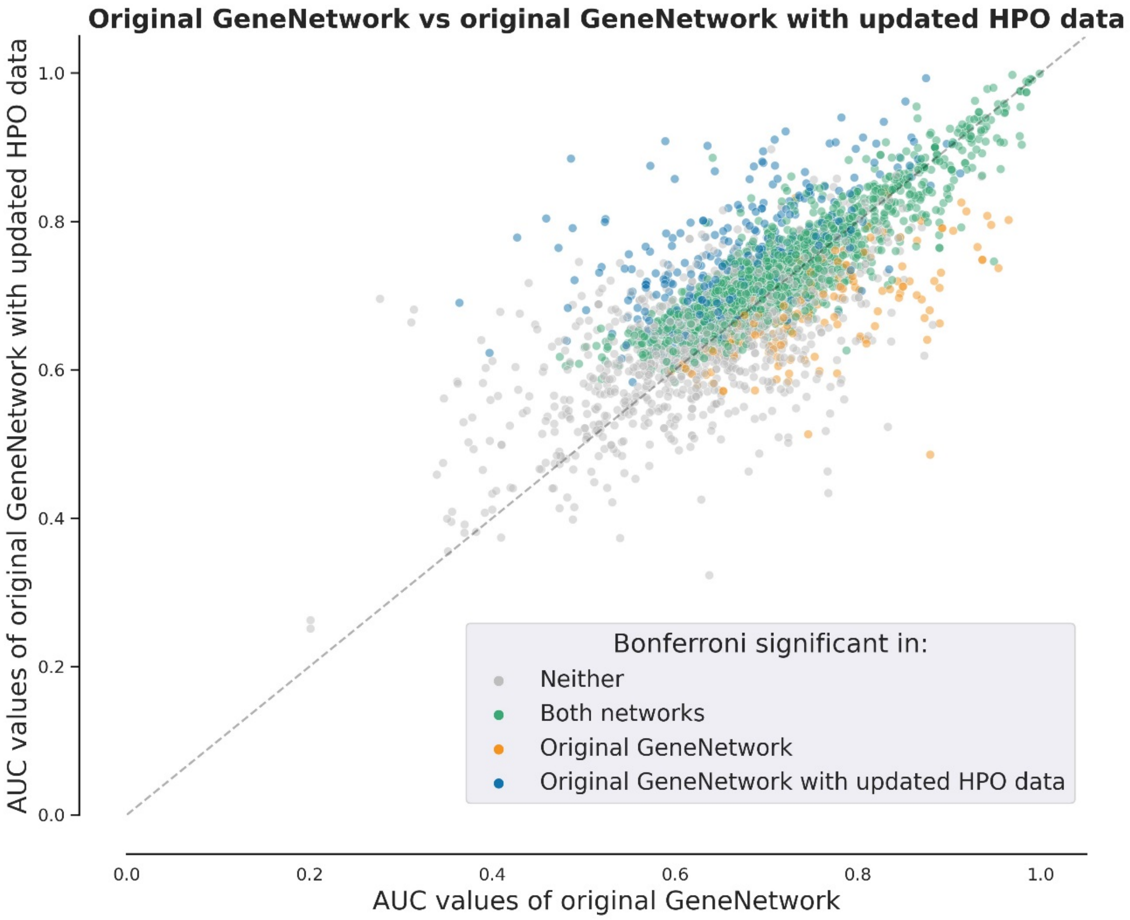

**Supplementary figure 7: AUC value comparison between the original GeneNetwork pipeline and the updated GeneNetwork pipeline with the original HPO data.** The prediction accuracy of the original GeneNetwork pipeline described in the GADO paper (x-axis) compared to the same pipeline with the updated HPO database described in this paper (y-axis). Every dot represents the AUC value of one HPO term. Dot color is based on the significance of the prediction in one, both, or neither methods after multiple testing correction. The mean AUC of the Original GeneNetwork is 0.69, and 1,751 pathways are predicted to be Bonferroni significant. The mean AUC of the updated GeneNetwork with original HPO data is 0.71, and 1,911 pathways are predicted to be Bonferroni significant.

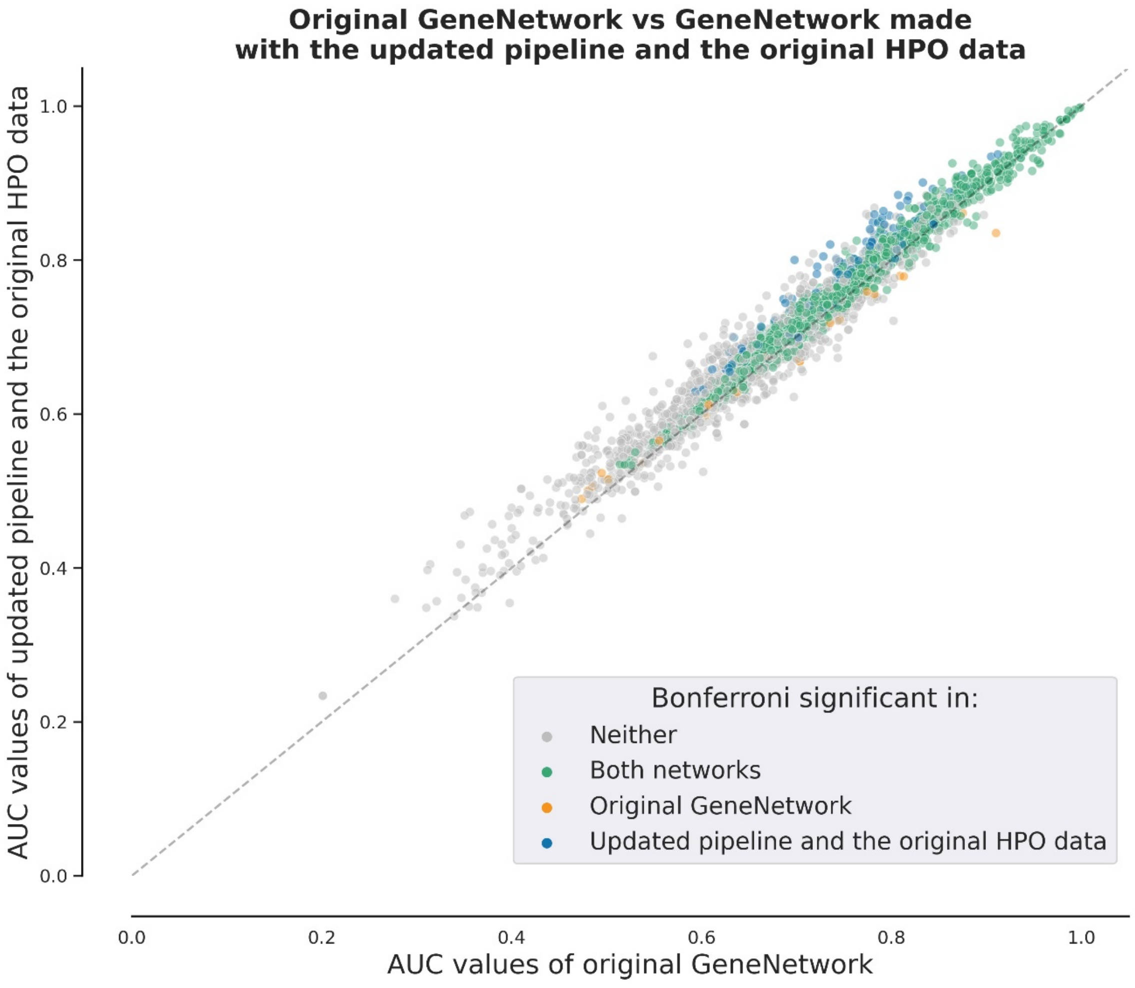

**Supplementary figure 8: AUC value comparison between the original GeneNetwork and the updated GeneNetwork.** The prediction accuracy of the original GeneNetwork pipeline described in the GADO paper (x-axis) is compared to the updated GeneNetwork (y-axis). Every dot represents the AUC value of one HPO term. Dot color is based on the significance of the prediction in one, both, or neither method after multiple testing correction. The mean AUC of the Original GeneNetwork is 0.69, and 1,751 pathways are predicted to be Bonferroni significant. The mean AUC of the updated GeneNetwork is 0.72, and 2,266 pathways are predicted to be Bonferroni significant.

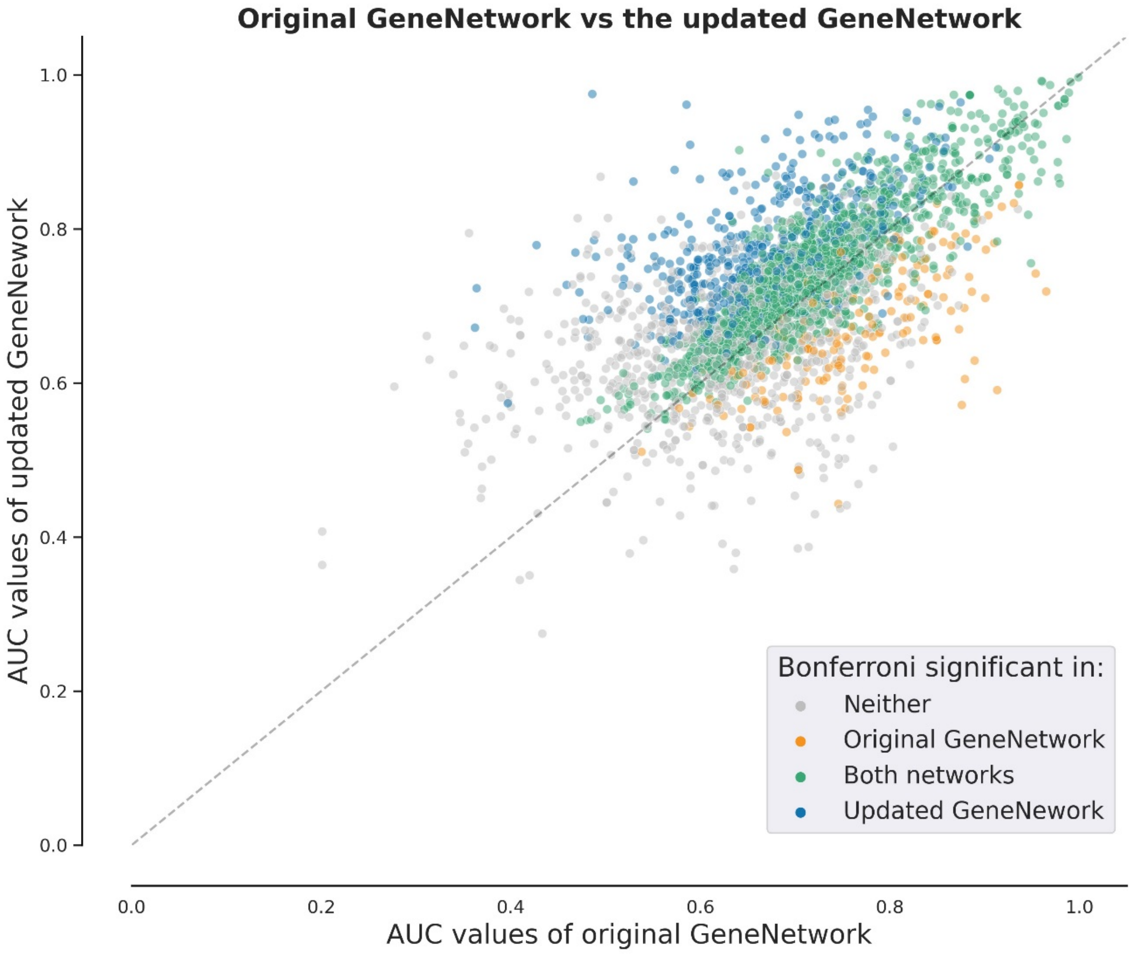

**Supplementary figure 9: AUC value comparison of kidney-related HPO terms between GeneNetwork and the network created with kidney-derived data.** Kidney-related HPO terms are predicted with a lower prediction accuracy compared to GeneNetwork. Furthermore, GeneNetwork is capable of predicting more kidney-related phenotypes compared to the network based solely on kidney samples.

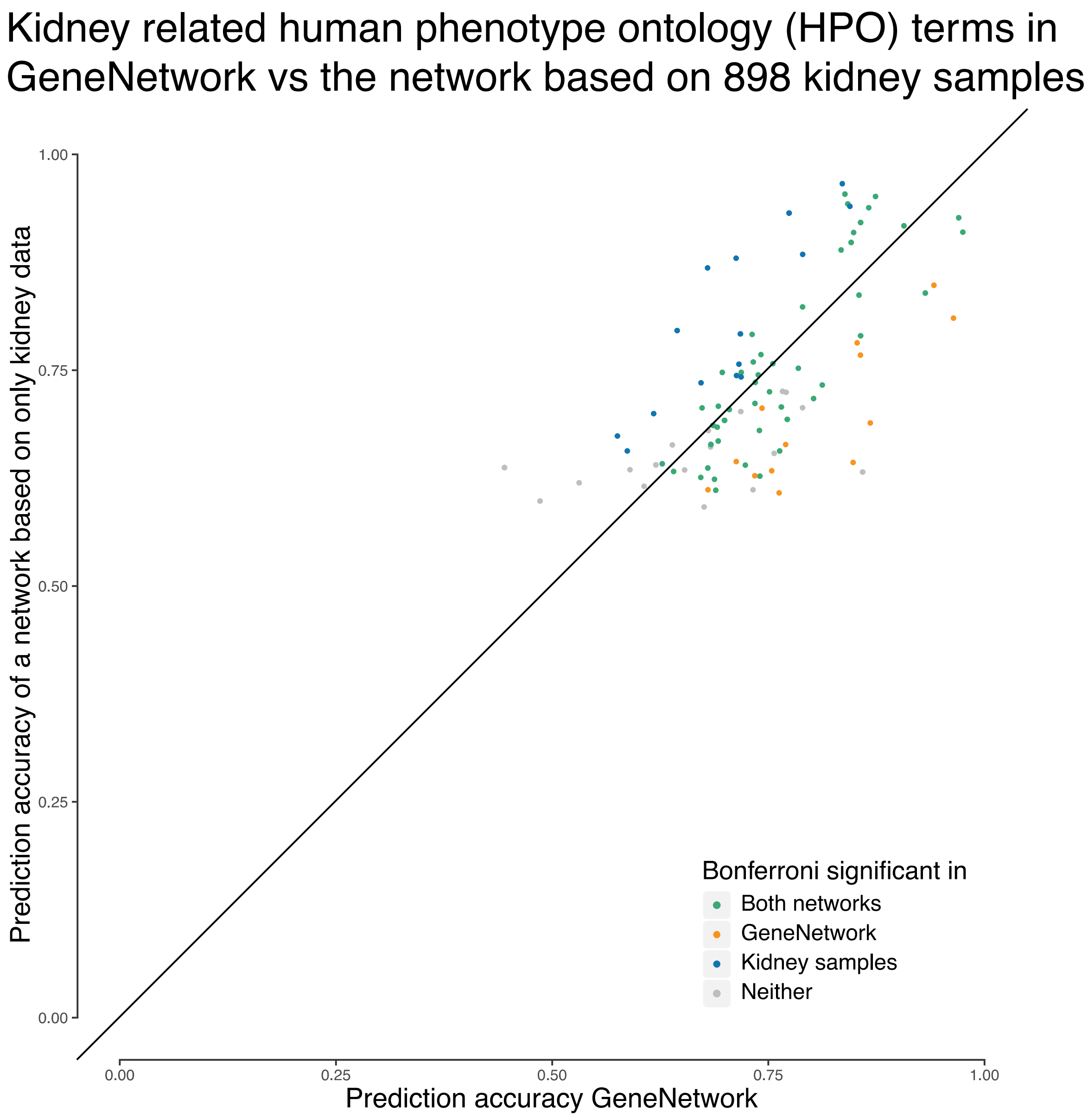

**Supplementary figure 10: *ALG6*-splice site analysis c.257+5G>A.** RNA of the index patient LE1 was extracted from whole blood and submitted to RT-PCR. Obtained cDNA was subsequently amplified for *ALG6* exons 3-5. In the patient LE1, two bands one wild type (WT) and one lower band could be detected upon electrophoresis (A). The lower band was cut from the gel and subsequently submitted for direct sequencing. Chromatogram revealed an in-frame deletion of ALG6-exon 4 (p.del57_86) (B-C).

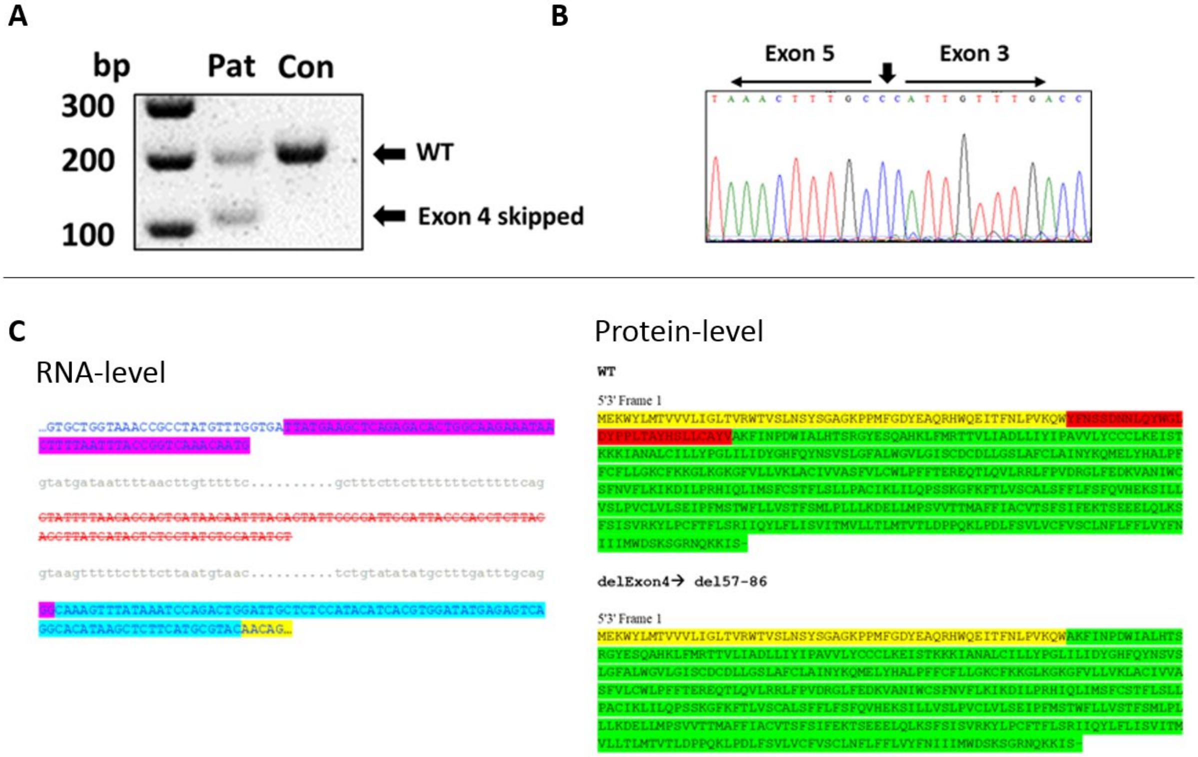

**Supplementary table 1. Keywords for kidney sample selection**

Keywords used for selection of kidney-derived RNA sequencing samples

*Supplied separately*

**Supplementary table 2. Sample annotations**

Description of the kidney samples and the original studies.

*Supplied separately*

**Supplementary table 3. Kidney-specific HPO terms**

Overview of kidney-specific HPO terms. We included HPO terms specific to the kidney for benchmark analysis. We excluded HPO terms related to kidney neoplasm and non-kidney specific child terms.

*Supplied separately*

**Supplementary table 4. Diagnostic kidney gene panel (n=379) of the University Medical Centre Utrecht NEF00v18.1**

Genes that were checked for (likely) pathogenic variants that might be causing the phenotype of patients with an *ALG6* variant.

*Supplied separately*

**Supplementary table 5. Improved kidney-specific HPO terms**

List of improved kidney-specific HPO terms in KidneyNetwork as compared to GeneNetwork

*Supplied separately*

**Supplementary table 6. Phenotypes and genes in 13 kidney disease patients**

Overview of phenotypes of the 13 patients that were analysed. For each patient, we identified which genes prioritized by GADO with KidneyNetwork overlap with genes containing potentially pathogenic variants predicted by CAPICE. The resulting gene lists contained 1–4 candidate genes for 9 of the 13 patients.

*Supplied separately*

**Supplementary table 7. Information on patients with *ALG6* variants deemed non-causative, including variant details and in silico predictions**

Overview of patients with *ALG6* variants deemed non-causative. The various reasons for exclusion are listed in the last column.

*Supplied separately*

**Supplementary table 8. Additional prediction scores for *ALG6* variant in patient GEL1**

Additional prediction scores for the variant found in patient GEL1: SIFT, PolyPhen-2, MutationTaster, Grantham Distance.

*Supplied separately*
